# Developmental mis-specification of cardiac conduction and structure transcriptome in Brugada syndrome

**DOI:** 10.64898/2026.07.20.26358469

**Authors:** Thomas Stervinou, Bastien Cimarosti, Robin Canac, Aurore Girardeau, Loubna Ahmed, Agnès Tessier, Jeremie Poschmann, Richard Redon, Flavien Charpentier, Patricia Lemarchand, Nathalie Gaborit, Guillaume Lamirault

## Abstract

**Background and Aims:** Brugada syndrome (BrS) is an inherited arrhythmia associated with ventricular fibrillation and sudden cardiac death. While pathogenic *SCN5A* variants account for approximately 20-25% of cases, most patients have no identifiable causal rare variant. Genome-wide association studies have identified common susceptibility variants near transcription factor genes involved in cardiac development, suggesting that developmental abnormalities may contribute to BrS pathogenesis beyond sodium channel dysfunction. We therefore investigated whether genetically distinct forms of BrS exhibit altered developmental transcriptomic trajectories during human cardiogenesis.

**Methods:** Daily bulk 3′ RNA sequencing was performed throughout a 30-day directed cardiac differentiation of hiPSC lines derived from two healthy controls and four BrS patients representing distinct genetic backgrounds: *SCN5A* haploinsufficiency (BrS-SCN5A and BrS-SCN5A-2), a pathogenic *RRAD* variant (BrS-RRAD), and a patient without rare pathogenic variants but carrying a high burden of common BrS susceptibility alleles (ACV).

**Results:** Transcriptomic trajectories markedly differed according to the underlying genetic architecture. While the BrS-SCN5A line remained largely similar to controls throughout differentiation, BrS-RRAD and BrS-ACV lines diverged as early as day 5, corresponding to the onset of cardiac specification. Differential expression analysis identified only 113 dysregulated genes in the BrS-SCN5A line compared with 525 and 383 genes in the BrS-RRAD and BrS-ACV lines, respectively. The BrS-RRAD and BrS-ACV models shared a common developmental signature characterized by early downregulation of key regulators of cardiac conduction system development, including *NKX2-5*, *IRX3* and *IRX5*, together with upregulation of genes involved in extracellular matrix organization. Interestingly, similarly to the BrS-SCN5A line, the BrS-SCN5A-2 line presented a transcriptomic remodeling that diverge from the BrS-RRAD and BrS-ACV lines. Consistent with these findings, predicted IRX5 regulatory normal interactions were completely lost in both non-*SCN5A* models, whereas transcriptomic remodeling remained minimal in both the BrS-SCN5A hiPSC line and an independent *Scn5a* haploinsufficient mouse model.

**Conclusion:** BrS associated with *SCN5A* haploinsufficiency and non-*SCN5A* genetic backgrounds follows distinct developmental transcriptomic trajectories. Our findings support a model in which non-*SCN5A* BrS is associated with early dysregulation of developmental transcriptional networks controlling cardiac conduction and extracellular matrix organization, whereas *SCN5A*-mediated BrS primarily results from sodium channel deficiency with limited developmental remodeling. These results identify developmental heterogeneity as a potential determinant of BrS pathophysiology and clinical variability.

## Introduction

Brugada syndrome (BrS) is an inherited arrhythmia syndrome characterized by ST-segment elevation in the right precordial leads and an increased risk of ventricular fibrillation and sudden cardiac death^1,2^. Since the identification of *SCN5A* as the first disease-causing gene, BrS has been widely regarded as the archetypal inherited cardiac channelopathy^3,4^. Approximately 20-25% of patients carry pathogenic loss-of-function variants in *SCN5A* gene, leading to reduced cardiac sodium current (I_Na_) and impaired electrical conduction^5^. In addition, rare variants in a limited number of other genes, including *RRAD* gene, account for only a small proportion of cases^6^. Collectively, currently recognized rare genetic variants explain less than one third of BrS patients, indicating that additional mechanisms contribute to disease susceptibility and suggesting that BrS is genetically and biologically more heterogeneous than initially appreciated.

Several independent observations now challenge the concept of BrS as a disorder resulting exclusively from ion-channel dysfunction. Genome-wide association studies have consistently identified common susceptibility variants located predominantly within non-coding regions surrounding genes encoding transcription factors involved in early cardiac development, including regulators of ventricular specification and maturation, and cardiac conduction system formation^7–9^. In parallel, increasing clinical and experimental evidence has revealed subtle structural abnormalities in subsets of BrS patients, including myocardial fibrosis and localized conduction slowing, particularly within the right ventricular outflow tract^10–12^. Together, these findings raise the possibility that, at least in part, BrS may originate from subtle disturbances of cardiac development that subsequently create an arrhythmogenic substrate^13,14^.

Testing this hypothesis directly in humans has remained challenging. Analyses performed in adult myocardial tissue cannot readily distinguish primary developmental abnormalities from secondary remodeling associated with aging, recurrent arrhythmias or disease progression^15,16^. Human induced pluripotent stem cell-derived cardiomyocytes (hiPSC-CMs), however, recapitulate the sequential stages of human cardiogenesis and therefore provide a unique opportunity to investigate developmental mechanisms underlying inherited cardiac diseases. Longitudinal transcriptomic profiling throughout directed cardiac differentiation allows reconstruction of developmental trajectories and identification of the earliest molecular events that may predispose to disease before the acquisition of mature cardiomyocyte phenotypes^17^.

Whether distinct genetic forms of BrS present and share common developmental mechanisms remains unknown. In particular, it is unclear whether *SCN5A*-related BrS and non-*SCN5A* BrS arise through similar biological pathways or instead reflect fundamentally different pathogenic processes^6,18,19^. Resolving this question is essential to understand the remarkable clinical variability of BrS and may help explain why patients carrying different genetic backgrounds frequently exhibit distinct electrophysiological phenotypes and disease severity.

We therefore hypothesized that BrS is associated with early transcriptomic alterations during human cardiogenesis and that these developmental trajectories differ according to the underlying genetic architecture. To test this hypothesis, we performed daily bulk 3′ RNA sequencing throughout a 30-day directed cardiac differentiation of hiPSC lines derived from control individuals and three genetically distinct BrS patients: one carrying a pathogenic *SCN5A* loss-of-function variant, one carrying a pathogenic *RRAD* variant, and one carrying no rare pathogenic variant but a high burden of common BrS susceptibility alleles. By reconstructing developmental transcriptomic trajectories, we sought to determine when disease-associated alterations first emerge and whether developmental remodeling differs between various genetic forms of BrS.

## Methods

### Human induced pluripotent stem cells (hiPSC) reprogramming and maintenance

All cell lines used in this paper have been already characterized. The 2 control lines Ctl-1 (IRX-5-Wt)^20^ and Ctl-2 (WT8288)^21^ as well as BrS-SCN5A (BrS2^+^)^22^, BrS-SCN5A-2 (BrS1^+^)^22^ and BrS-RRAD (BrS5^-^)^22,23^ have been generated using Sendai virus method. BrS-ACV (BrS6^-^)^22,24^ has been generated using lentiviral method. All the hiPSC lines were maintained at 37°C, 5% CO_2_, 21% O_2_ in StemMACS^TM^ iPS Brew WF Medium (Miltenyi Biotec, Bergisch Gladbach, Germany) on culture plates coated with Matrigel^®^ hESC-Qualified Matrix (0.05 mg/mL, Corning, NY, USA). At 75% confluency, the cells were passaged using Gentle Cell Dissociation Reagent (STEMCELL^TM^ Technologies, Vancouver, Canada).

### Cardiac Differentiation of hiPSCs

The directed cardiac differentiations of the hiPSCs were performed using the established matrix sandwich method^17^. When the hiPSCs reached 90% confluency, an overlay of Growth Factor Reduced Matrigel (0.033 mg/mL, Corning) was added. Differentiation was initiated 24 h later by culturing the cells in RPMI1640 medium (Thermo Fisher Scientific, Waltham, MA, USA) supplemented with B27 (without insulin, Thermo Fisher Scientific), 2 mM L-glutamine (Thermo Fisher Scientific), 1% NEAA (Thermo Fisher Scientific), 100 ng/mL Activin A (Miltenyi Biotec), 1X Pen/Strep (Thermo Fisher Scientific) and 10 ng/mL FGF2 for 24 h. Subsequently, on the next day, the medium was replaced by RPMI1640 medium supplemented with B27 without insulin, 2 mM L-glutamine, 1% NEAA, 10 ng/mL BMP4 (Miltenyi Biotec), 1X Pen/Strep and 5 ng/mL FGF2 for 4 days. By day 5, cells were cultured in RPMI1640 medium supplemented with B27 complete (Thermo Fisher Scientific), 2 mM L-glutamine, 1X Pen/Strep and 1% NEAA and changed every two days until day 30.

### Bulk Transcriptomics

#### RNA extraction and sequencing

For each hiPSC line, the samples were harvested daily, from D-1 to D30 of the cardiac differentiation protocol, from three independent cardiac differentiations. The total RNA were extracted using the NucleoSpin RNA kit (MACHEREY-NAGEL, Hoerdt, France) and their quality was assessed by NanoDropTM 1000 Spectrophotometer (Thermo Fisher Scientific). From the D-1 to D14 samples, all of the cells were collected while, from D15 to D30, to obtain samples enriched with cardiomyocytes, only the spontaneously beating cell clusters were collected, following mechanical isolation using a needle. Three RNA libraries were prepared by GenoBiRD core facility according to their published method^25^and sequenced on 8 individual runs on a NovaSeq 6000 or HiSeq 2500 Sequencing System (Illumina, San Diego, CA, USA).

#### Primary Analysis of Bulk Transcriptomic Data

Demultiplexing, alignment on the GRCh38 reference genome and counting steps were conducted on each sequencing run with the Snakemake pipeline developed by the GenoBiRD core facility^25^. Normalized and log-transformed expression matrices were generated using the multiplates function correcting potential batch effects by treating cardiac differentiation time points as replicates.

#### PCA

Principal component analysis (PCA) were performed using the R package FactoMineR^26^ on the entire mean-centered and log-transformed matrix. Trajectories trends were implemented with weighted regression smoothing in the ggplot2 R package^27^ using geom_smooth() function.

#### Time-Course Differentially Expressed Genes Analysis

Significantly deregulated genes between both control lines (Ctl-1 and Ctl-2) and each BrS line during time-course directed differentiation were identified using a robust longitudinal composite method of three different approaches (RegROTS, DiffROTS and PolyROTS) with the R package RolDE^28^, applied on the log-transformed matrix of the selected samples. Genes with pvalue adjusted <0.05 have been selected.

#### Differentially Expressed Genes Analysis

Significantly deregulated genes at specific time points were identified using the R package DESeq2^29^. Counts were normalized and processed by the R package Combat-seq^30^. Genes with p-value adjusted < 0.05 were selected as differentially expressed.

#### Heatmap

Differentially expressed genes of each comparison were grouped into clusters, based on their expression level variation across the directed cardiac differentiation, using the k-means method, and were visualized with the ComplexHeatmap R package^31^.

#### Gene Ontology enrichment analysis

Gene Ontology (GO) analysis were performed using the R package ClusterProfiler^32^ based on GO Biological Process terms. Significantly enriched (bonferroni-corrected p-value < 0.05) biological processes, as compared to reference transcriptome, with a Gene Set Size (GSSize) between 10 and 500, and a minimum of 3 genes per functions, were considered for further analysis. The 15 GO terms with the lowest corrected p-values were selected and visualized with treeplot.

#### Predicted interactions of transcription factors

For each hiPSC line, the gene regulatory network was inferred using the LEAP (Lag-based Expression Association for Pseudotime-series) R package^33^, based on the average of the log-transform values from three independent cardiac differentiations. Cardiac differentiation time points were used to rank samples, as required, by the LEAP tool. The max_lag_prop parameter was set to 1/10, meaning that, calculation of the maximum absolute correlation (MAC) score was performed on a 3-day window, at most. Only gene-to-gene associations with a significant MAC score (permutation test p-value < 0.05) and with a non-null time window, were considered. Gene-to-gene associations with a positive correlation score were interpreted as activation relationships and those with a negative correlation score as repression relationships.

## Results

### Patient characteristics and genetics

Three patients affected by type I BrS (BrS-SCN5A, BrS-RRAD and BrS-ACV) with a familial history of SCD or syncope were selected (Figure 1A). Patient #1, BrS-SCN5A, was an asymptomatic 55-year-old male whose brother died suddenly at age 38, previously described^22^. He presented a spontaneous BrS ECG pattern with occurrence of VF during an electrophysiological study. An ICD was implanted with no occurrence of syncope or VF during a 13-year follow-up. Six of his relatives presented with a BrS ECG pattern including his son who exhibited one spontaneous episode of VF. Patient #2, BrS-RRAD, was a 41-year-old male, previously described^22,23^, who presented recurrent near syncopes with palpitations and spontaneous BrS ECG pattern. Electrophysiological study induced VF. An ICD was implanted with no recurrence of syncope or VF during a 16-year follow-up. Familial screening identified 6 relatives with a BrS ECG pattern and one with unexplained SCD at age 41. Patient #3, BrS-ACV, was a 42-year-old male, previously described^22,24^. While he had a spontaneous BrS ECG pattern, he presented an unexplained syncope, and suffered from an out-of-hospital cardiac arrest at night. Three of his relatives presented with a BrS phenotype after ajmaline administration.

**Figure 1.**
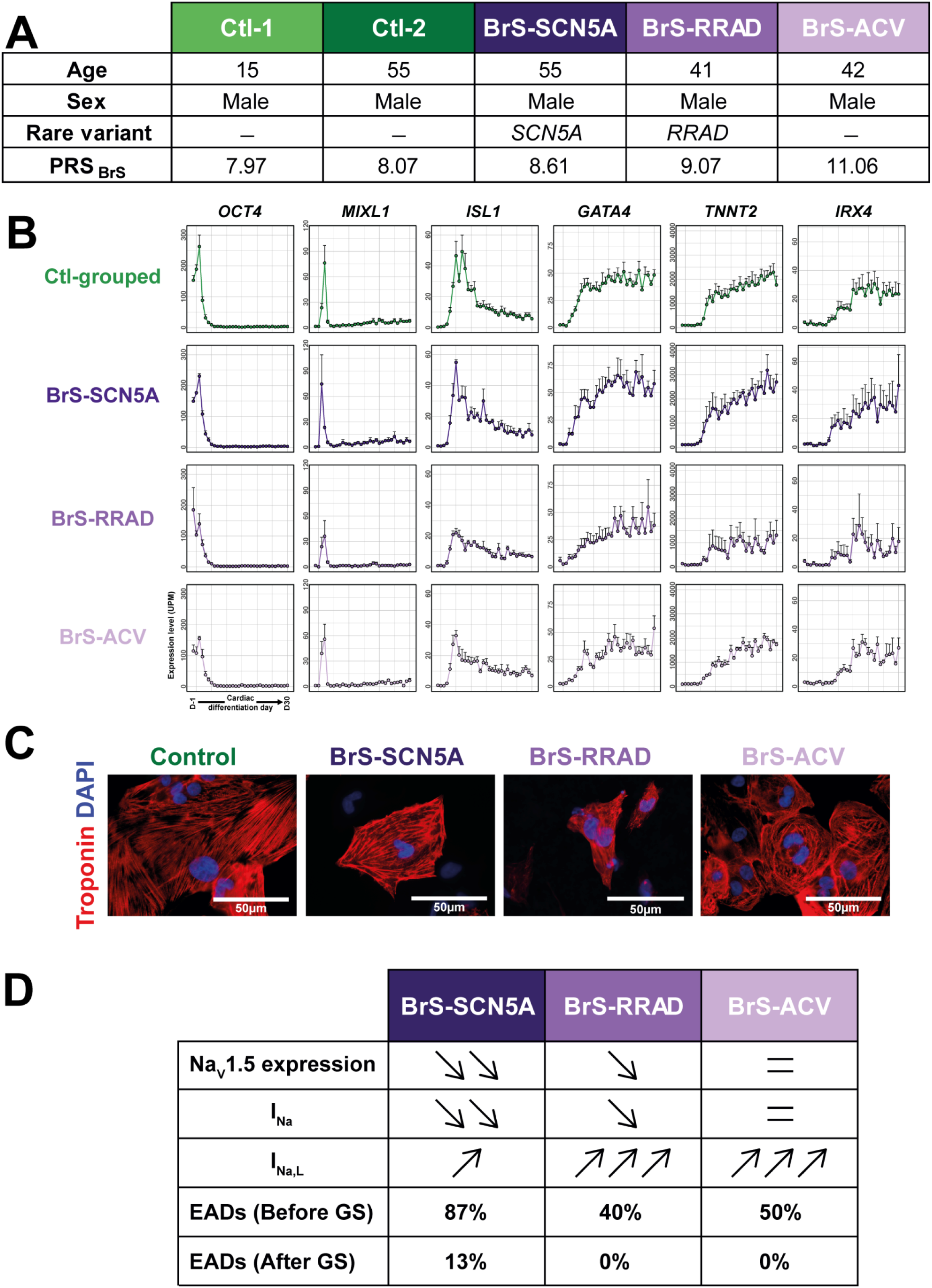
Control and Brugada patient hiPSCs lines genotyping and phenotyping. (A) Table showing the genotype and phenotype of control individuals and BrS patients. (B) Expression of cardiac developmental-specific markers at each day of cardiac differentiation: OCT4, pluripotency; MIXL1, primitive mesoderm; ISL1, cardiac progenitors; GATA4 and TNNT2, cardiomyocytes; IRX4, ventricular identity. (C) Immunofluorescence of troponin in cardiomyocytes differentiated from each hiPSC line. (D) Electrophysiological characteristics of BrS lines. Control lines exhibited 4% EADs before GS (not shown) [22]. PRS: Polygenic risk score; GS: GS-458967

Two control male subjects were also included (Figure 1A). One subject who was an unrelated healthy extra familial control (Ctl-1) and the other (Ctl-2) was an unaffected relative of BrS-RRAD patient, in whom BrS was excluded after sodium channel blocker challenge^20–22^. At the genetic level, screening of rare variants in the coding regions of 20 genes, selected according to the American College of Medical Genetics and Genomics (ACMG) guidelines, was performed for all 5 lines. It identified an *SCN5A* pathogenic rare variant in BrS-SCN5A patient. He carried a 10bp duplication (c.1983–1993dup) in *SCN5A* creating a stop codon (p.A665G-fsX16). This variant has been functionally investigated, showing that it leads to haploinsufficiency of *SCN5A*^22^. BrS-RRAD patient presented a pathogenic rare genetic variant in the *RRAD* gene (p.R211H) whereas no rare genetic variant was identified in BrS-ACV patient, nor in Ctl lines^22–24^

Genome-wide association studies (GWAS) were also performed in all 5 lines to investigate presence of common susceptibility variants associated to BrS, in order to calculate, based on 21 risk alleles and their corresponding effect sizes, their respective polygenic risk score (PRS_BrS_)^8^. Interestingly, BrS-ACV displayed the highest PRS_BrS_ (11.06), whereas the PRS_BrS_ for Ctl and both rare variant-carriers (BrS-SCN5A and BrS-RRAD) were homogeneously much lower (Figure 1A).

### Generation and characterization of hiPSC cardiac differentiations

Somatic cells were obtained from each studied individual and were reprogrammed into hiPSC lines, as previously described^22^. We investigated whether the process of cardiac differentiation could be altered in BrS lines. We generated daily transcriptomic data, from the hiPSC stage (D-1) to day 30 (D30), for three independent cardiac differentiations of each of the five hiPSC lines. The kinetic expression of key markers of each stage of the cardiac development process, was similar among all lines, showing that all BrS hiPSC lines properly followed the expected transcriptomic variations associated to the directed cardiac differentiation (Figure 1B). At the end stage of the directed cardiac differentiation, cells expressed cardiac-specific troponin I, demonstrating their capability to form functional CMs (Figure 1B,C). Previous electrophysiological examination of these hiPSC-CMs showed that the 3 BrS lines presented variations in their end-stage phenotype. The expression of Nav1.5 and the density of its corresponding I_Na_ current was strongly reduced in the BrS-SCN5A CMs, while it was mildly and not reduced in BrS-RRAD and BrS-ACV CMs, respectively. Inversely, the late sodium current (I_Na,L_) was mildly increased in BrS-SCN5A CMs, while it was strongly increased in BrS-RRAD and BrS-ACV CMs. Overall, the I_Na,L_ to I_Na_ peak current ratio was strongly increased in all 3 lines as compared to control with the same order of magnitude^22^. All lines showed arrhythmogenic phenotype marked by early after depolarizations, whereas EADs were absent from control lines. However, although BrS-SCN5A CMs showed the highest amount of EADs, they could not be fully suppressed by the use of I_Na,L_ inhibitor (GS-458967), in opposition to BrS-RRAD and BrS-ACV (Figure 1D). Overall, this shows that the directed cardiac differentiation process gives rise to CMs with comparable maturity level for all 5 lines; that the BrS lines display an expected sodium current alteration and proarrhythmogenic phenotype; and suggests that BrS-RRAD and BrS-ACV share a closer electrophysiological phenotype, as compared to the BrS-SCN5A.

### Transcriptomic signatures associated to each BrS lines in hiPSC-CMs

We then investigated whether these BrS CM phenotypic differences were underpinned by alterations at the transcriptomic level. First, we investigated the specific transcriptomic alterations in each BrS-lines, performing a differential gene expression analysis of each BrS line as compare to the Ctl lines, during the course of the directed differentiation protocol. We found that, as compared to the BrS-RRAD (n=525) and BrS-ACV lines (n=383), BrS-SCN5A line presented with the lowest number of differentially expressed genes (n=113; Figure 2A). Accordingly, ventricular transcriptomic analysis of a murine model of BrS, carrying heterozygote deletion of *SCN5A,* also showed very limited remodeling (Supplemental figure 1)^22^. This result has also been observed in another murine model of *SCN5A* haploinsufficiency^23^. The misregulated gene-clusters in BrS-RRAD and BrS-ACV lines were associated to cardiac function and contraction, but also, in a large extend, to developmental processes (Supplemental figure 2). Similarly, the few biological functions associated to BrS-SCN5A line were also linked to developmental processes but in a much lower extend. Accordingly, the number of differentially expressed genes associated to these developmental processes was lower in BrS-SCN5A line (n=14 genes, 12.4% of DEGs) as compared to the 2 other lines (BrS-RRAD : n=148, 28.1% of DEGs; BrS-ACV: n=95; 24.8% of DEGs; Supplemental figure 2).

**Figure 2.**
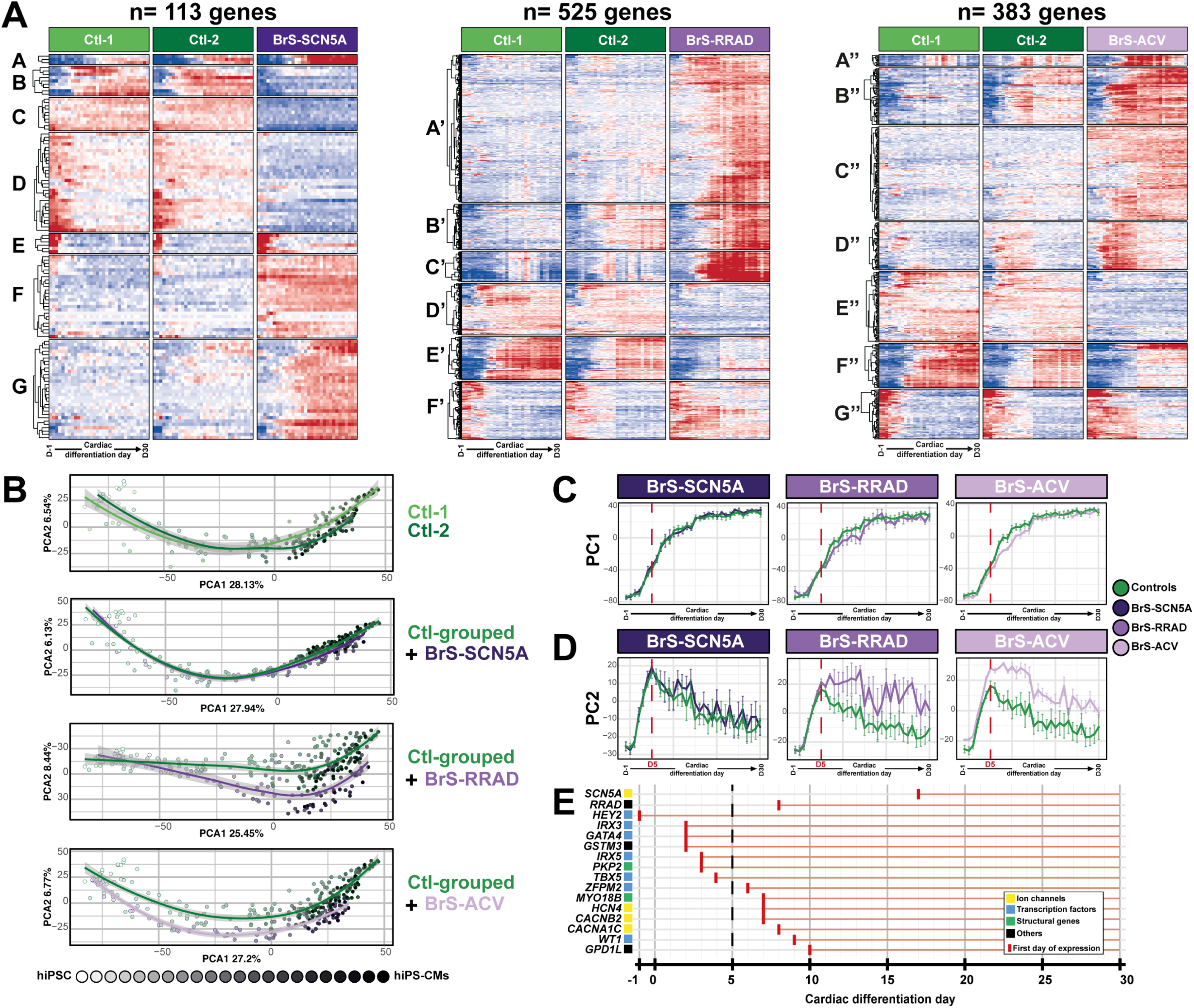
Transcriptomic remodeling of BrS lines over the course of cardiac differentiation. (A) Heatmap of differentially expressed genes over the course of cardiac differentiation. Each row represents a gene. Each column represents a specific time point of differentiation. For each cell line, columns are ordered according from hiPSC stage to cardiac stage, from left to right. Red represents higher expression and blue, lower expression level. In each cluster, genes are grouped in clusters according to their proximity in expression pattern. (B) Principal component analysis (PCA) of the directed cardiac differentiation for each line. Each data point is filled with a color ranging from white, for hiPSC stage, to black, for hiPS-CMs stage. (C-D) Values of the principal component 1 (C) or 2 (D) over the course of the differentiation. (E) Expression profile of major genes associated to BrS genetic diagnosis in control lines. Red vertical bars and horizontal lines represent the onset, and the maintenance of expression, respectively, for each of these genes during the cardiac differentiation process.

To explore this developmental link to BrS, we investigated BrS transcriptomic alterations at the kinetic level. We performed PCA analyses, using all expressed genes (n=24305 genes). Transcriptomic profile from the two Ctl lines did not significantly differ (Figure 2B). Regarding BrS vs Ctl comparisons, PCA1, showed similar temporal evolutions for all comparisons (Figure 2B,C). In opposition, PCA2 displayed variabilities between some of the lines: while BrS-SCN5A transcriptomic trajectory was close to the Ctl lines, the BrS-RRAD and BrS-ACV, significantly diverge from the Ctl lines and from the BrS-SCN5A line transcriptomic trajectory (Figure 2B,D). More precisely, we found that the transcriptomic divergence, seen in PCA2, of BrS-RRAD and BrS-ACV as compared to Ctl lines, started early during the differentiation, at day 5 (Figure 2D). This differentiation stage corresponds to early cardiac developmental stage in this model^17^. Interestingly, the expression of the majority of BrS-reported culprit genes onsets around day 5 (Figure 2E). All in all, this showed that BrS-RRAD and BrS-ACV have both stronger transcriptomics alterations as compared to BrS-SCN5A line, starting early in the developmental process (Day 5).

### Transcriptomic signatures associated to each BrS line

Using the transcriptomic dataset, generated in kinetics, we identified genes that were commonly or specifically altered in each BrS line as compared to Ctl. We identified 64, 393 and 266 genes upregulated in respectively, BrS-SCN5A, BrS-RRAD and BrS-ACV lines (Figure 3A), and 49, 132 and 117 downregulated genes (Figure 3D). We grouped them in clusters according to their expression profile, showing variable temporal expression patterns in both up and down regulated genes. Among upregulated genes, 8 genes were altered in all 3 BrS lines, 11 were altered in both the BrS-SCN5A and BrS-RRAD lines, 23 in both the BrS-SCN5A and BrS-ACV lines and 97 in both the BrS-ACV and BrS-RRAD lines (Figure 3B). Among downregulated genes, 2 were altered in all 3 BrS lines, 5 were in both the BrS-SCN5A and BrS-RRAD lines, 12 in both the BrS-SCN5A and BrS-ACV lines and 41 in both the BrS-ACV and BrS-RRAD lines (Figure 3E). We then investigated whether these shared gene alterations were preferentially associated to specific clusters, and therefore to specific gene signatures. We found that, for both BrS-ACV and BrS-RRAD lines specifically, almost each cluster (all but one) in one line was preferentially linked to a single cluster in the other line (Figure 3C,D). This suggests that BrS-ACV and BrS-RRAD lines share specific biological function alterations. To further investigate this hypothesis, we specifically examined the expression patterns and biological functions of the 128 up or down regulated genes in both BrS-ACV and BrS-RRAD lines, but not in BrS-SCN5A line, as compared to Ctl lines. They were grouped in 6 clusters, each with specific expression pattern (Figure 4A, Supplemental Figure 3). In accordance with the altered propagation of the electrical influx in Brugada-affected patients, mis-regulated genes in both BrS-ACV and BrS-RRAD lines were associated, in part, to functional and structural processes, such as ion homeostasis and extracellular matrix organization (Figure 4B). Interestingly, other functional annotations were related to developmental processes (Figure 4B). Accordingly, these gene clusters mostly showed transcriptional mis-regulation starting early during the differentiation process in BrS-ACV and BrS-RRAD lines and that were maintained until the end of the protocol (day 30). Interestingly, these clusters included a set of 13 transcription factors, four being downregulated, *IRX2*, *IRX3*, *IRX5* and *NKX2-5*, and the remaining nine TFs being upregulated (Figure 4C, 4D, Supplemental Figure 4). These misregulations were absent in BrS-SCN5A line (Figure 4C, 4D, Supplemental Figure 4).

**Figure 3.**
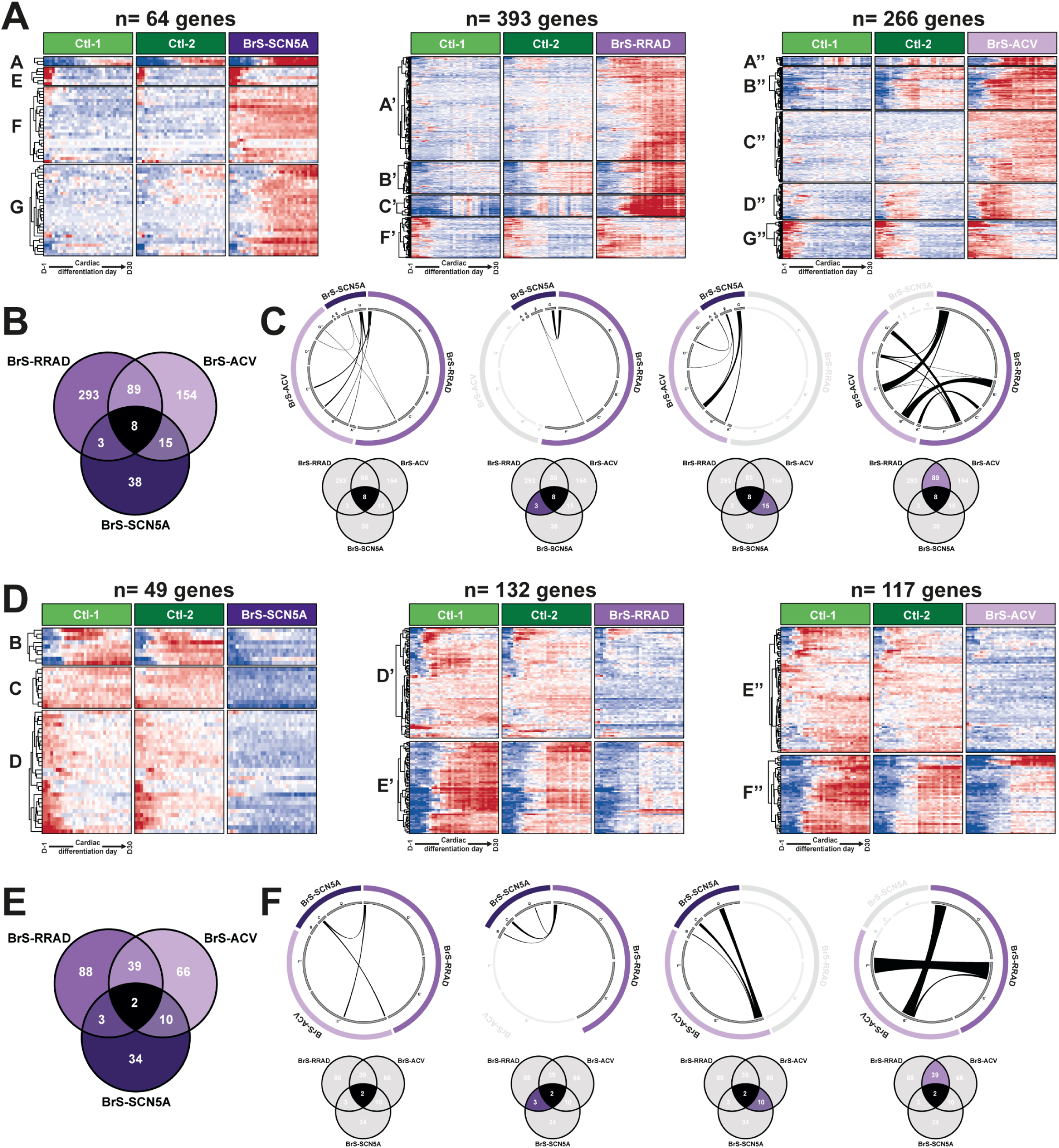
Comparison of up and down regulated genes in each BrS line. (A-C) Up-regulated genes. (D-F) Down-regulated genes. Heatmaps of (A) up-regulated and (D) down-regulated genes in each BrS line as compared to control. Venn Diagram of (B) up-regulated and (E) down-regulated genes in each BrS line as compared to control. (C,F) ChordPlot of the (C) up-regulated and (F) down-regulated genes shared between conditions. Each arc represents how a gene connects between cluster in the compared conditions.

**Figure 4.**
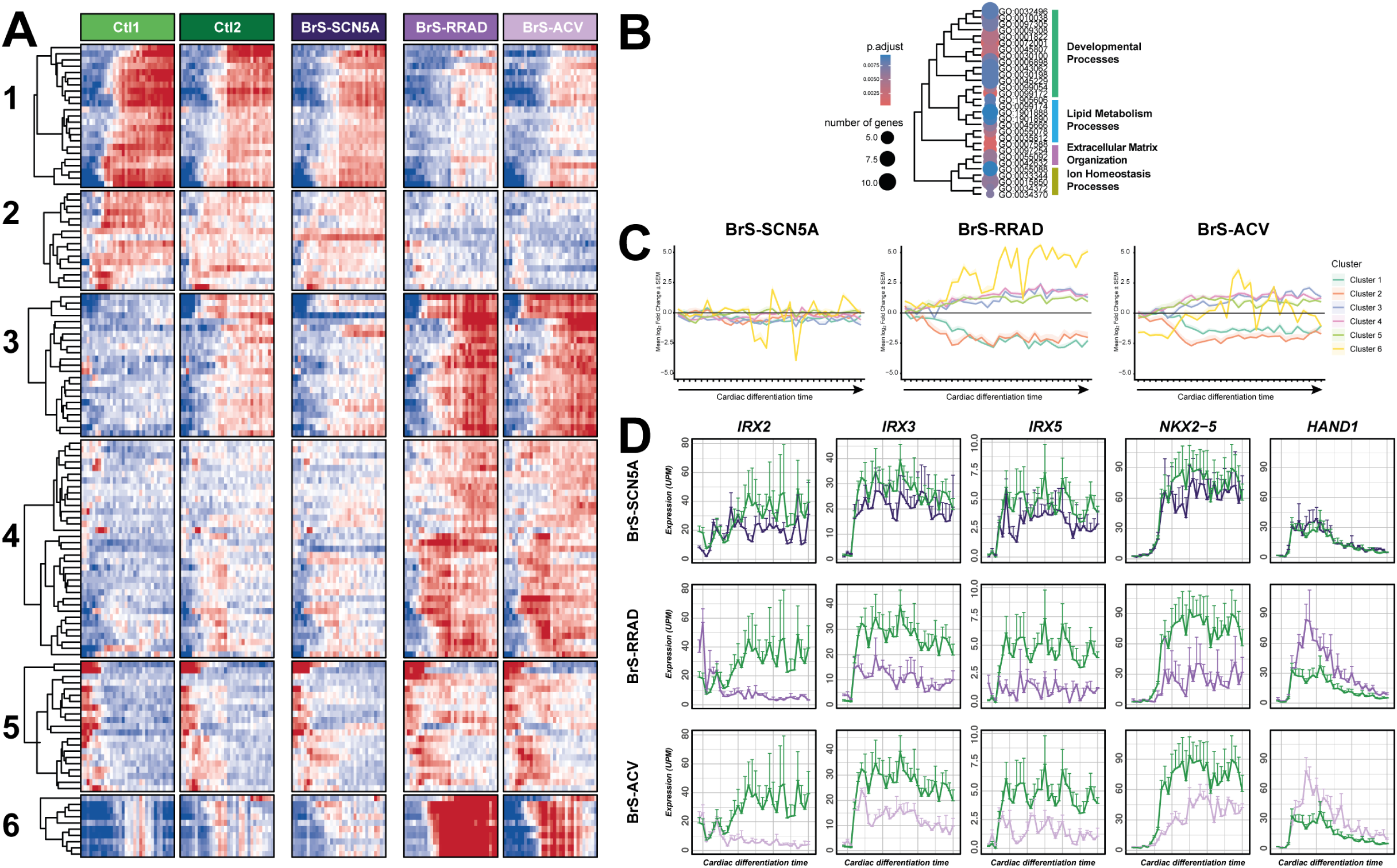
Transcriptomic signature common to BrS-RRAD and BrS-ACV lines. (A) Heatmap of the 128 differentially expressed genes in both BrS-RRAD and BrS-ACV lines. (B) Gene enrichment analysis of this 128-gene signature. The size of the dot point represents the number of genes and the color represents the corresponding adjusted p-value. (C) Mean log-fold change for each of the 6 clusters over the course of the directed cardiac differentiation. (D) Expression in Units Per Million (UPM) of a selection of five out of the 13 cardiac transcription factors (IRX2, IRX3, IRX5, NKX2-5, and HAND1) which expression is altered in both BrS-RRAD and BrS-ACV lines during the directed cardiac differentiation.

To confirm that this transcriptional misregulations were consistently absent in the setting of BrS with *SCN5A* mutation, we also investigated this signature in the BrS-SCN5A-2 line, at day 30 of the cardiac differentiation. Interestingly, the expression pattern obtained for 2 independent clone of the BrS-SCN5A-2 line was comparable to the one of BrS-SCN5A line and clearly diverged from the ones of BrS-ACV and BrS-RRAD lines (Figure 5A,B).

**Figure 5.**
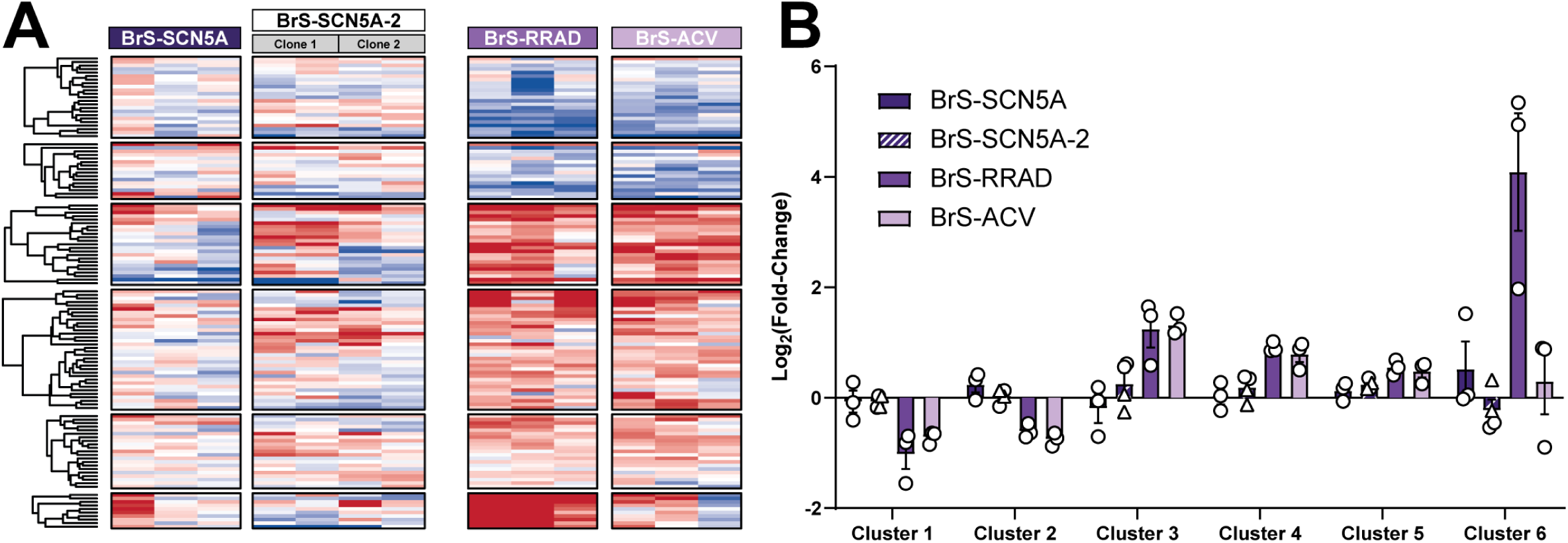
Comparison of the BrS-RRAD / BrS-ACV 128-gene signature in SCN5A-mutated and non SCN5A-mutated BrS lines. (A) Expression level heatmap of the 128 genes in 2 SCN5A-mutated lines: BrS-SCN5A and BrS-SCN5A-2, as compared to the non SCN5A-mutated lines. Expression level for two independent clones are displayed for BrS-SCN5A-2. (B) Bar graph displaying the average Log2 fold change of expression for each line as compared to control. Data is presented cluster by cluster.

### Transcriptomic regulations specific to each BrS line

We then aimed at further defining the potential role of these 13 TFs in BrS-associated transcriptomic alterations. We first unveiled their putative target genes over the course of the directed cardiac differentiation in control cells within the 128-gene BrS-RRAD/BrS-ACV signature. Most of the identified targets genes were putatively regulated by several of the 13 TFs, with the main pattern being a combination of activation and repression effect (Figure 6A). Interestingly, two of these TFs, IRX3 and IRX5, displayed highly comparable target-gene lists with comparable regulatory effects (activation / repression), with 82.1% and 74.2% common activations in IRX3 and IRX5 respectively, and 90.1% common repressions in IRX3 and IRX5 (Figure 6A, B). This finding is consistent with the fact that they share high level of expression pattern similarity across all tissues and organs, as well as combinatorial roles in cardiac structure and function regulation^36,37^.

**Figure 6.**
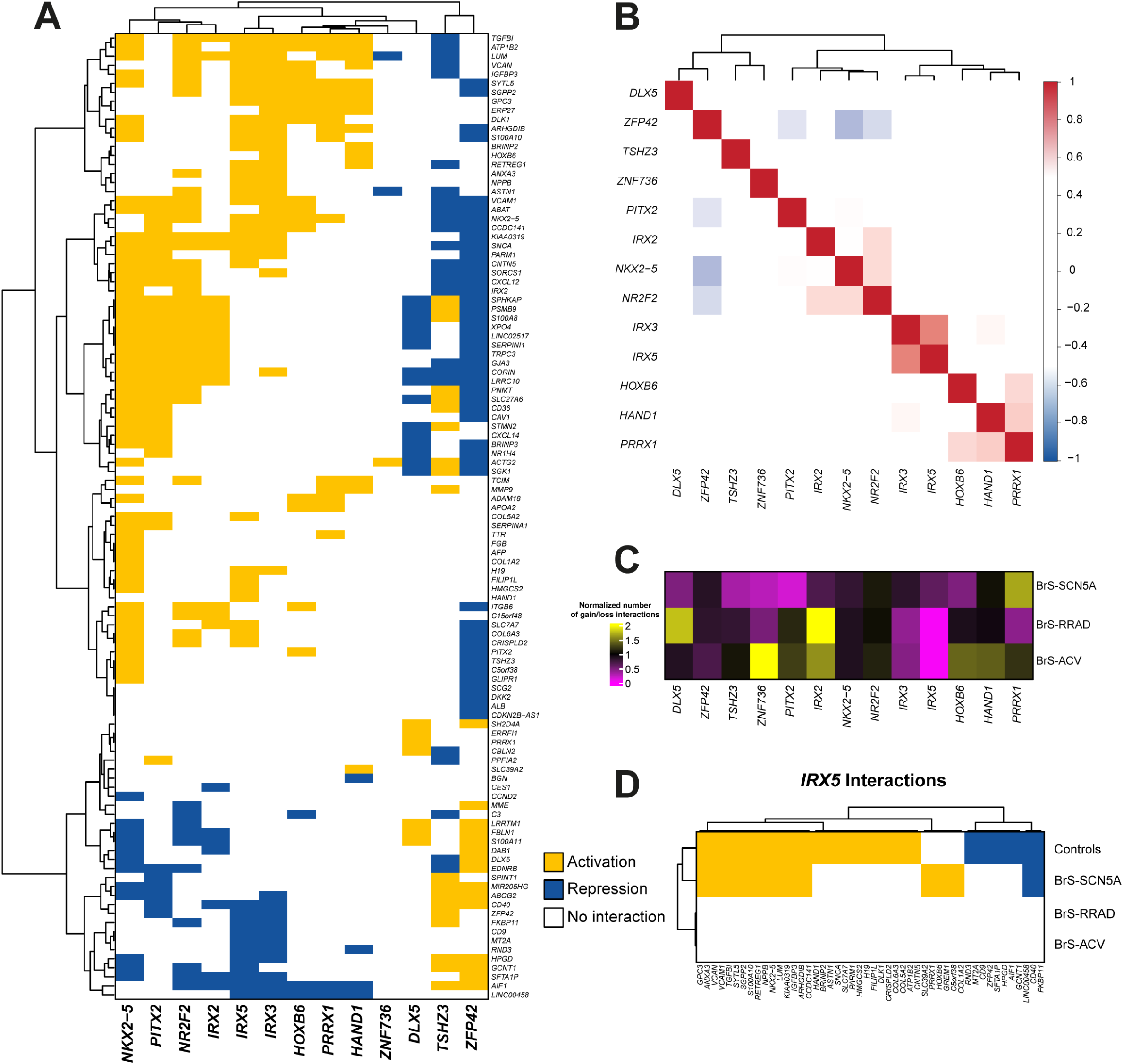
Predictions of activation and repression of the 128-gene signature by the 13 transcription factors . (A) Interactions prediction in control lines. Yellow color represented an activation, blue color, a repression of expression, and white, absence of interaction. Each column represent a transcription factor and each line represents the genes predicted to display at least one activation or repression. (B) Correlation matrix of the predicted interaction patterns among all 13 transcription factors. Red color represents a positive correlation and blue color, a negative correlation. (C) Normalized number of interactions (gain and loss) in BrS-SCN5A, BrS-RRAD and BrS-ACV lines, as compared to controls, for each transcription factor. Yellow color represents a higher number of interactions and pink, a lower number of interactions. (D) Interactions prediction for IRX5 transcription factor in all lines for genes that displayed at least one activation or repression in at least one line. Yellow color represented an activation, blue color, a repression of expression, and white, absence of interaction.

We then investigated if the interactions between each TF and their target genes were altered in the context of BrS. Activation and repression effects for each BrS cell line were identified and compared to the results obtained in the control cell line. Gain and loss of activation / repression effects in each BrS cell line, as compared to the control line, are summarized in Figure 6C. Interestingly, the most striking finding was linked to IRX5 with 100% loss its interactions in BrS-RRAD and BrS-ACV lines as compared to control (Figure 6C). Figure 6D displays the IRX5 regulatory effect for control and BrS lines on the genes that were defined as an IRX5 target in at least one cell line. We found that in BrS-SCN5A line 21 activations were different from control, with 15 loss of activation and 6 gain of activation. Also 8 repressions were loss but no gain was observed. In strong opposition, BrS-RRAD and BrS-ACV displayed a loss of all their interactions, whether they were activation or repression, with no gain of interaction, as compared to Control (Figure 6D). No such striking findings were observed for the other 12 TFs (Supplemental Figure 5). These findings were coherent with previous hypothesis on the participation of IRX5 to BrS pathophysiology^38,42^.

### Participation of IRX5 downregulation to the phenotype of non SCN5A-mutated BrS lines

We further investigated the combinatorial role of IRX5 downregulation of expression and loss of regulatory interactions, observed in BrS-RRAD and BrS-ACV lines. We used a IRX5^KO^ hiPSC line previously generated from Control 1 line^20^, and confirmed the absence of IRX5 expression over the course of the directed cardiac differentiation (Figure 7A). We performed bulk transcriptomic analysis at day 12 of differentiation, corresponding to beginning of cardiac specification. This analysis revealed that 1553 genes were mis-expressed as compared to Control. At day 12, 525, 383 and 113 genes were differentially expressed in, respectively, BrS-RRAD, BrS-ACV and BrS-SCN5A, as compared to Control (Figure 7C). Interestingly, 98 (19%) and 117 (31%) of these genes were also misexpressed in the IRX5^KO^ line, for BrS-RRAD, BrS-ACV, while only 11 (10%) were also misexpressed in the IRX5^KO^ line for BrS-SCN5A line (Figure 7C). Figure 7D displays the expression profiles of the 98, 117 and 11 genes previously identified for all cell lines. Interestingly, regarding the list of 98 genes commonly misexpressed in IRX5^KO^ and BrS-RRAD, their expression pattern was very similar in IRX5^KO^, BrS-RRAD and BrS-ACV, but not in BrS-SCN5A. Likewise, expression pattern of the 117 genes commonly misexpressed in IRX5^KO^ and BrS-ACV lines, was very similar in IRX5^KO^, BrS-RRAD and BrS-ACV, but not in BrS-SCN5A. In opposition such finding was not observed for the 11 genes commonly misexpressed in IRX5^KO^ and BrS-SCN5A lines (Figure 7D).

**Figure 7.**
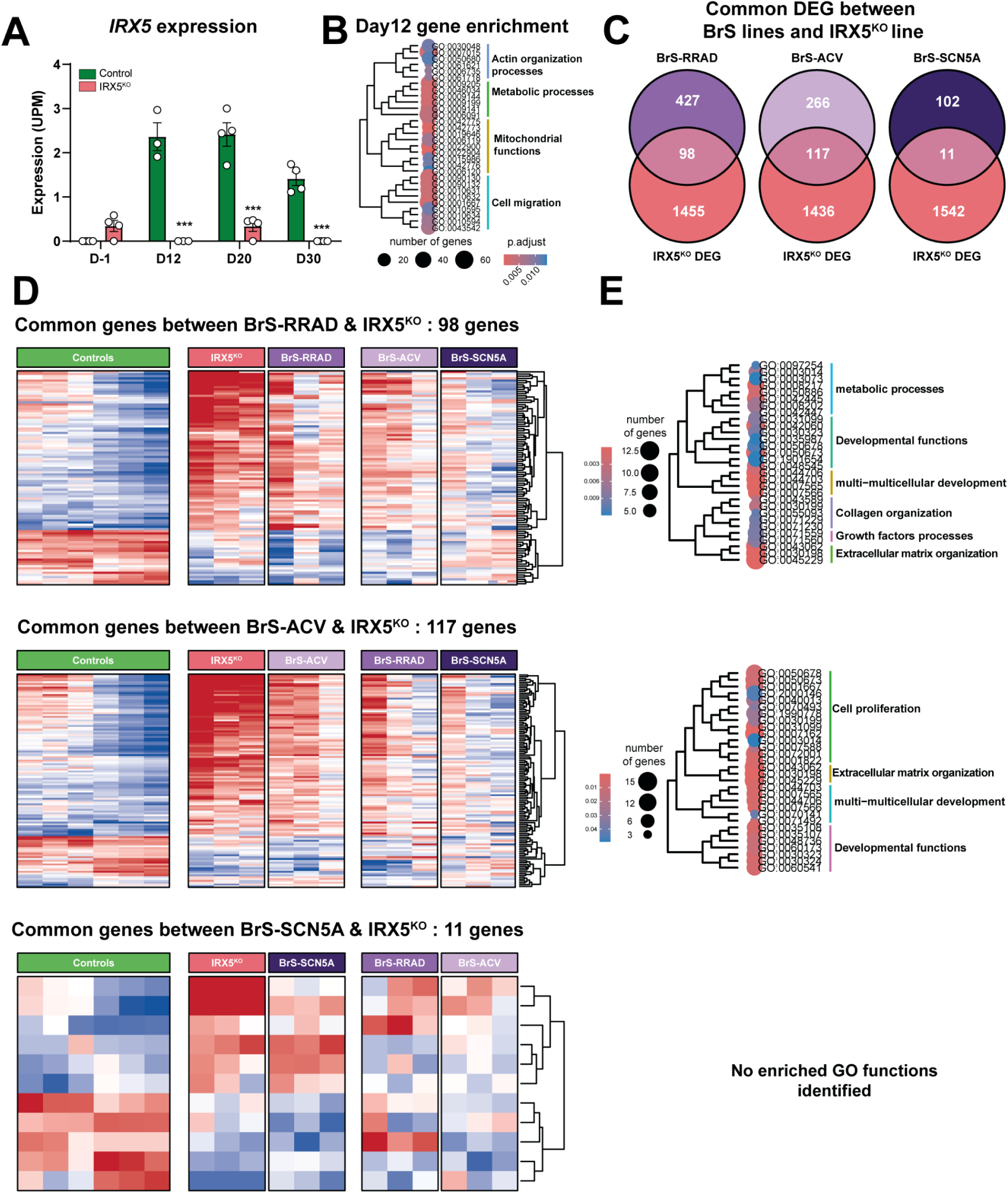
Comparison of transcriptomic remodeling between each BrS line and the IRX5^KO^ line. (A) Expression of IRX5 in control line and in the corresponding isogenic IRX5^KO^ at 4 time points of the cardiac differentiation, Day -1, hiPSC stage; Day 12, onset of cellular beating; Day 20 and Day 30, early and late cardiac maturation stages. (B) Gene enrichment analysis of the genes differentially expressed genes at day 12 of the cardiac differentiation, in IRX5^KO^ line as compared to Control line. The size of the dot point represents the number of genes and the color represents the corresponding adjusted p-value. (C) Venn Diagramm of genes altered in IRX5^KO^ and in each of the BrS line at day 12. (D) Heatmaps of the common altered genes in each BrS line as compared to IRX5^KO^. (E) Gene enrichment analysis of the genes differentially expressed genes in each heatmap. The size of the dot point represents the number of genes and the color represents the corresponding adjusted p-value. KO: Knock-out.

Gene ontology analysis of the 98 and 117 commonly altered genes with BrS-RRAD and BrS-ACV, revealed overrepresentation of biological processes associated to development and extracellular matrix organization (Figure 7E). In contrast, the gene ontology analysis of all IRX5^KO^ differentially expressed genes revealed overrepresentation of actin organization, metabolic processes and cell migration functions, but no biological process associated to development (Figure 7B). Similarly, the gene ontology analysis of the 11 commonly altered genes with BrS-SCN5A did not point to biological process associated to development (Figure 7E).

## Discussion

In the present study, we reconstructed the transcriptomic trajectories, throughout cardiac differentiation of hiPSC-derived cardiomyocytes, from genetically distinct Brugada syndrome (BrS) patients. Three major findings emerged. First, BrS associated with RRAD mutation or with an accumulation of common susceptibility alleles displayed an early transcriptomic divergence from control cells beginning at the onset of cardiac specification, whereas the SCN5A-mutated line remained remarkably similar to controls. Second, the transcriptomic alterations observed in non-SCN5A BrS converged toward developmental pathways regulating cardiac conduction system specification and extracellular matrix organization. Third, these alterations were associated with reduced expression and profound disruption of the IRX5 regulatory network, supporting a role for altered developmental transcriptional programs in the pathophysiology of non-SCN5A BrS.

An important finding of our study is the marked contrast between SCN5A- and non-SCN5A-associated BrS. The SCN5A line exhibited only limited transcriptomic remodeling despite displaying a robust electrophysiological phenotype, consistent with our previous functional studies and with transcriptomic analyses from Scn5a haploinsufficient mice^34,35^. Together, these observations suggest that reduced N_aV_1.5 expression is a major driver of the BrS arrhythmogenic phenotype, without requiring extensive developmental remodeling. In contrast, both the RRAD and high polygenic risk lines shared widespread transcriptomic alterations affecting developmental transcription factors and structural pathways despite their distinct genetic backgrounds. These results support the concept that genetically distinct forms of BrS converge toward a similar clinical phenotype through different biological mechanisms^39^. Rather than representing a single disease mechanism, BrS may encompass at least two independent pathogenic trajectories: a primary sodium-channel disease driven by *SCN5A* haploinsufficiency and a developmental form characterized by coordinated dysregulation of transcriptional networks. How these trajectories combine or not with other disease mechanism such as auto-antibodies, remains to be explored^40,41^.

BrS has traditionally been considered the archetypal inherited cardiac channelopathy. However, this paradigm has progressively evolved following genome-wide association studies identifying susceptibility loci near transcription factor genes involved in cardiac development, including *TBX5*, *HEY2*, *NKX2-5* and members of the Iroquois, IRX, family^7,8^. While these studies suggested that developmental mechanisms may contribute to disease susceptibility, direct evidence in human cardiogenesis has been lacking. By following transcriptomic changes throughout cardiac differentiation, our study demonstrates that molecular divergence in non-*SCN5A* BrS begins as early as day 5 of differentiation, corresponding to early cardiac specification^17^. These alterations were maintained until the end of the differentiation, stage at which they were, once again, present in 2 non *SCN5A-*mutated lines with different genetic backgrounds, and absent in 2 *SCN5A*-mutated lines that both lead to haploinsufficiency of N_aV_1.5. These findings therefore support the hypothesis that developmental mis-specification contributes to BrS pathophysiology in non *SCN5A-*mutated BrS patients.

Among these developmental regulators, IRX5 emerged as a central candidate^38,42^. *IRX5* expression was selectively reduced in non-*SCN5A* BrS, its predicted regulatory interactions were almost completely lost, and a substantial proportion of dysregulated genes overlapped with those identified in IRX5-deficient hiPSC-cardiomyocytes. Moreover, these shared genes were enriched for biological processes related to cardiac development and extracellular matrix organization. Although these observations do not establish IRX5 as the primary causal mechanism, they identify disruption of the IRX transcriptional network as a plausible contributor to the developmental phenotype observed in non-*SCN5A* BrS. The concomitant downregulation of IRX2, IRX3 and NKX2-5 further suggests that impairment of a broader developmental regulatory network, rather than alteration of a single transcription factor, underlies these transcriptomic changes.

Our data also provide new insights into the increasingly recognized structural component of BrS. Histopathological and imaging studies have reported localized myocardial fibrosis, particularly within the right ventricular outflow tract, although its temporal origin remains uncertain^10,43–45^. We found early activation of transcriptional programs involved in extracellular matrix organization and collagen regulation during cardiomyocyte differentiation exclusively in non-SCN5A BrS lines. These findings do not demonstrate the development of fibrosis *per se* but suggest that the molecular programs predisposing to extracellular matrix remodeling are established during early cardiogenesis. This observation provides a potential developmental explanation for the subtle structural abnormalities increasingly reported in BrS patients.

Several limitations should be acknowledged. The study included a limited number of hiPSC line for each genetic mechanism, limiting the assessment of inter-individual variability within each subgroup. In addition, hiPSC-derived cardiomyocytes represent an immature developmental model and bulk RNA sequencing does not resolve cell type-specific transcriptomic changes. Finally, transcriptional interactions inferred by LEAP require further functional validation. Nevertheless, the remarkable convergence between two genetically distinct non-*SCN5A* BrS models, together with the limited remodeling observed in both *SCN5A* human and murine models, supports the robustness of our conclusions.

Overall, our findings suggest that developmental transcriptomic remodeling is a prominent feature of non-*SCN5A* BrS but not of *SCN5A*-mediated disease. These results support a model in which Brugada syndrome comprises biologically distinct pathogenic mechanisms that converge toward a common arrhythmogenic phenotype. Understanding this developmental heterogeneity may improve disease stratification and provide new opportunities for mechanism-based therapeutic approaches beyond sodium channel dysfunction.

## Data Availability

All data produced in the present study are available upon reasonable request to the authors

## Acknowledgments, and fundings

The authors are grateful to the patients and families who agreed to participate in our research. This work was supported by grants from the Fondation pour la Recherche Médicale (DEQ20140329545), National Research Agency ANR-14-CE10-0001-01 and La Fédération Française de Cardiologie. N.G. was laureate of grants from Fondation Lefoulon-Delalande and Marie Curie Actions, International Incoming Fellowship FP7-PEOPLE-2012-IIF (PIIF-GA-2012-331436) and from the National Research Agency (HEART-iPS ANR-15-CE14-0019-01). B.C. is laureate of fellowships from the Foundation pour la Recherche Médicale (FRM, PBR201810007614) and the Foundation Genavie. R.C. is laureate of fellowship from the Fonds Marion Elizabeth Brancher. B.C. and G.L. have been awarded a grant from The French Regional Council of Pays de la Loire (RFI project VaCaRMe).

## Supplemental data

**Supplemental figure 1.**
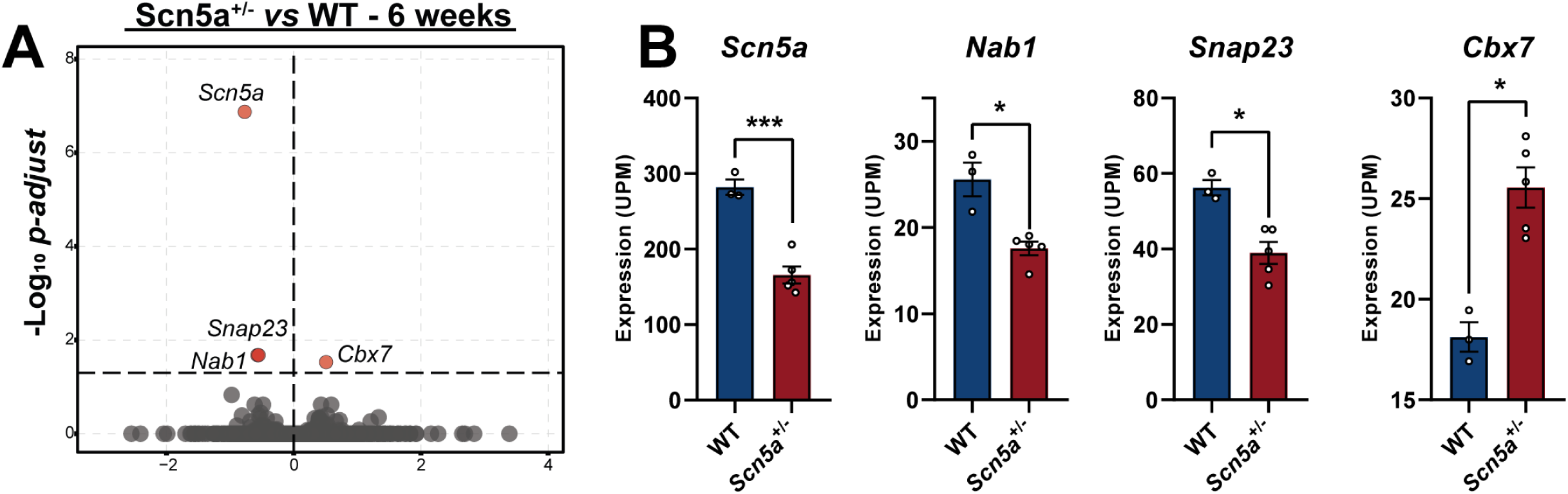
Transcriptomic analysis of ventricular samples of Scn5a^+/-^ murine model. (A) Volcano plot of gene expression in SCN5A^+/-^ as compared to WT murine model. Horizontal line represent the significance threshold defined as adjusted p-value < 0.05. (B) Bar graphs showing the expression levels of the 4 differentially expressed genes. * adjusted p-value <0.05 ; *** adjusted p-value <0.001.

**Supplemental figure 2.**
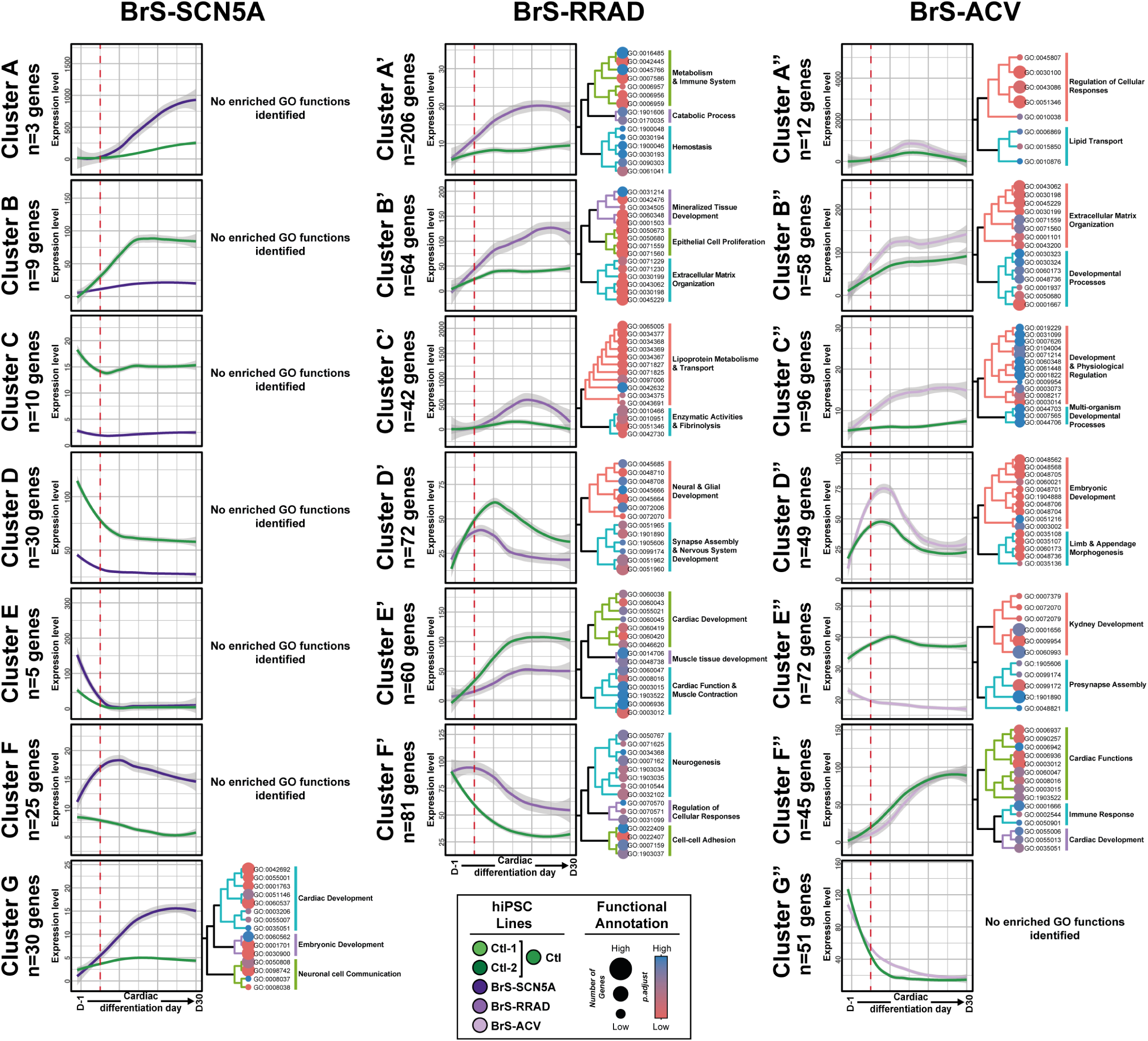
Expression pattern and associated overrepresented biological processes of all misregulated genes in each BrS line. For each BrS line, genes were grouped by clusters according to their expression patterns. The average gene expression of the genes within each cluster is represented in green for Control lines and in purple for BrS lines. Expression levels represent the values in Units per Million (UPM). Gene expression is ordered from right to left, from the hiPSC stage to the hiPS-CMs stage. Each cluster is associated to a treeplot of the top 15 overrepresented biological processes, when statistically significant.

**Supplemental figure 3.**
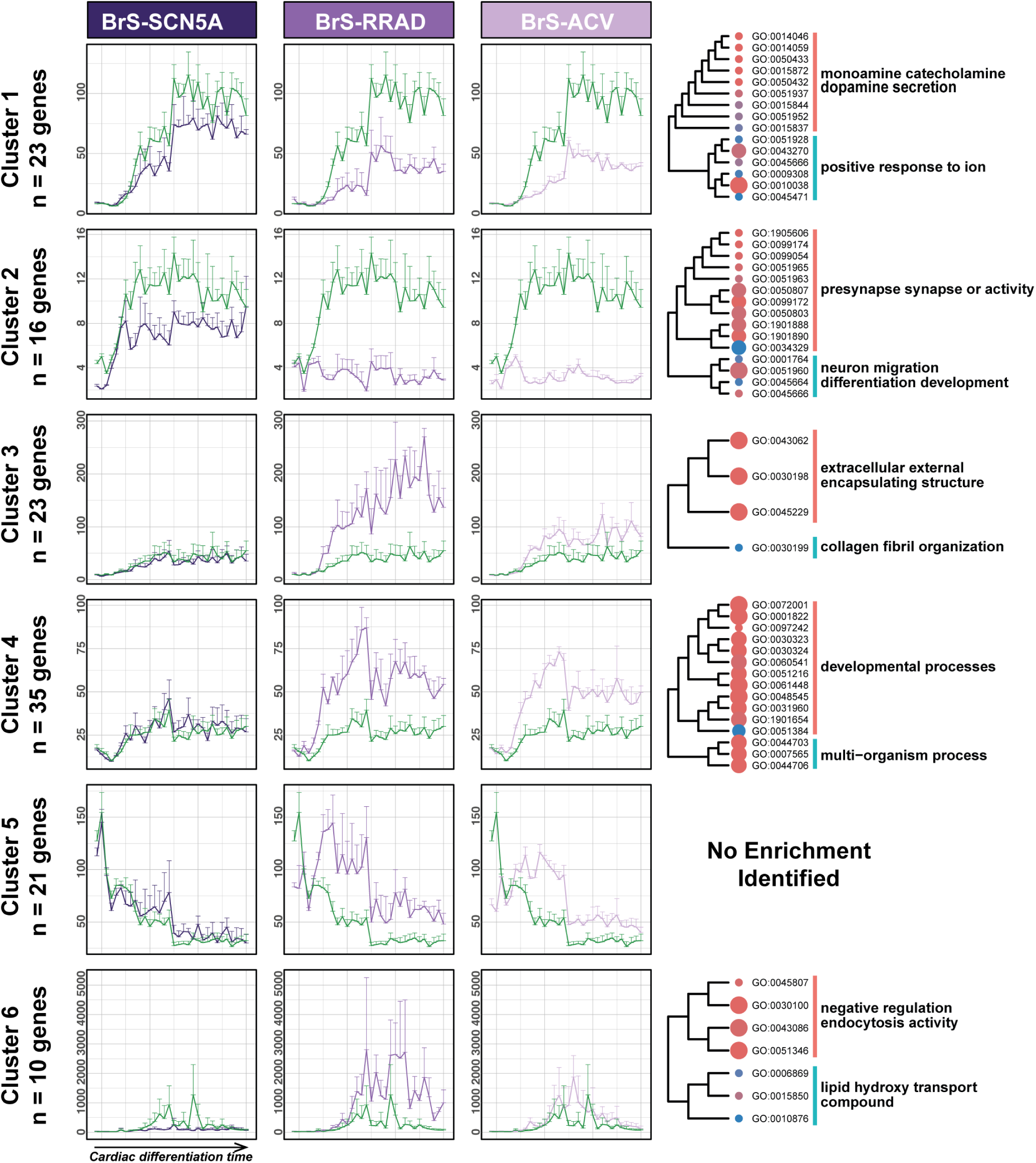
Expression pattern of the 128 BrS-RRAD/BrS-ACV gene signature and associated overrepresented biological processes, in the 3 BrS lines. For each BrS line, the average gene expression of the genes within each cluster is represented in green for Control lines and in purple for BrS lines. Expression levels represent the values in Units per Million (UPM). Gene expression is ordered from right to left, from the hiPSC stage to the hiPS-CMs stage. Each cluster is associated to a treeplot of the top 15 overrepresented biological processes, when statistically significant.

**Supplemental figure 4.**
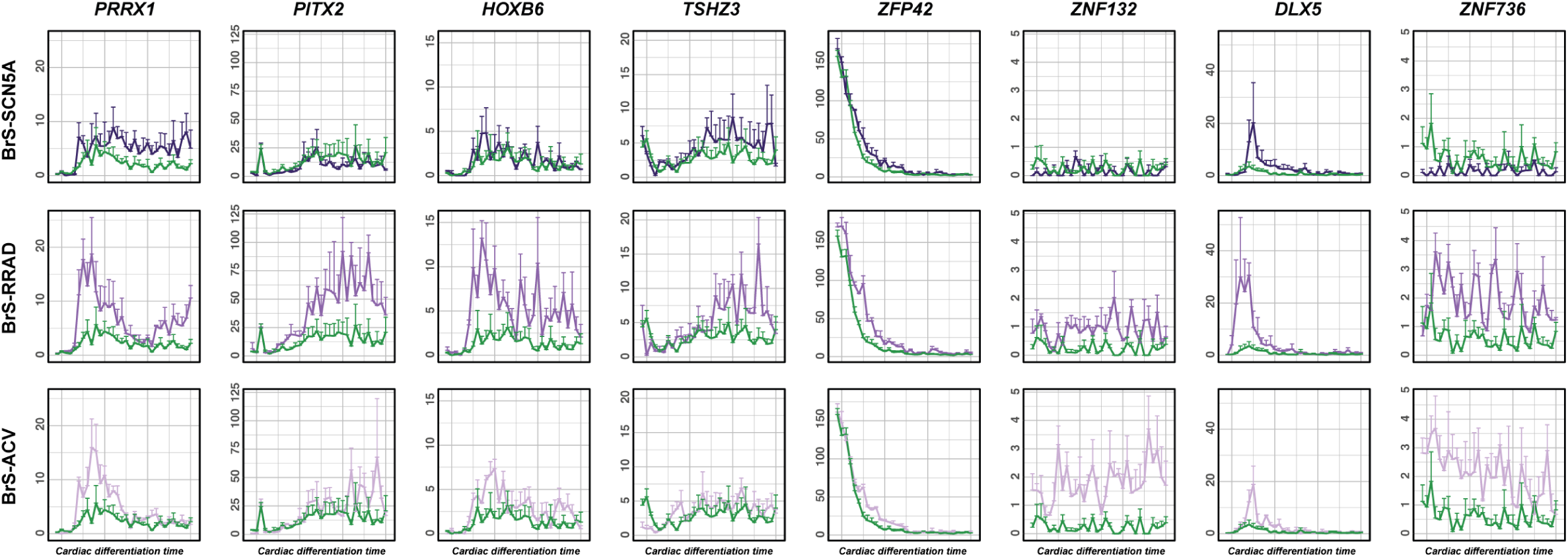
Expression of 8 transcription factors misregulated in BrS-RRAD and BrS-ACV lines. Each pattern of expression is displayed for each BrS line. Gene expression is ordered from right to left, from the hiPSC stage to the hiPS-CMs stage, respectively. The expression level is represented in green for Control lines and in purple for BrS lines. Expression levels represent the values in Units per Million (UPM).

**Supplemental figure 5.**
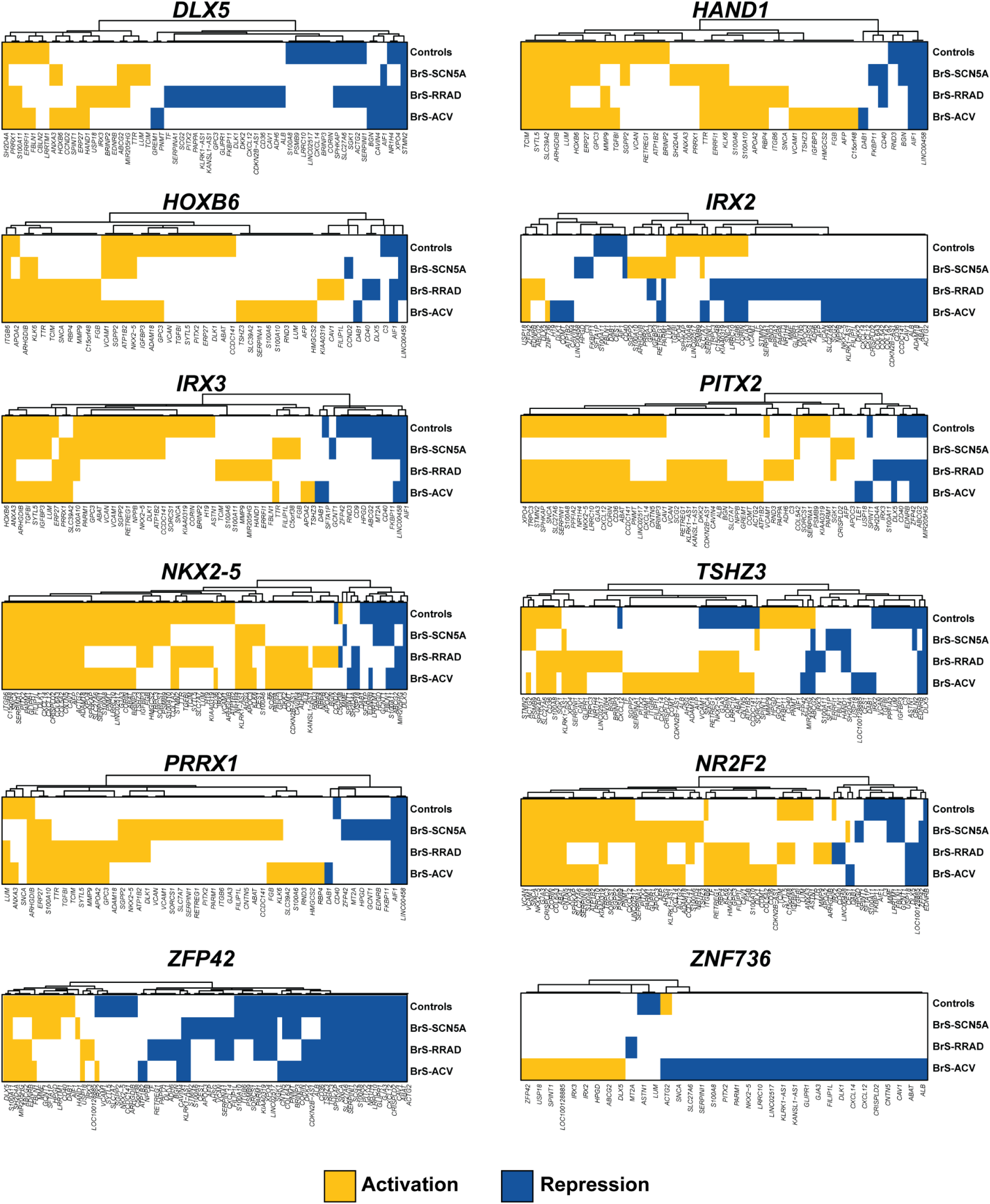
Prediction of interactions of 12 transcription factors of the common misregulated genes in BrS-RRAD/BrS-ACV signature. Interactions prediction in all lines for genes that displayed at least one activation or repression in at least one line. Yellow color represented an activation, blue color, a repression of expression, and white, absence of interaction.

